# Fully automatic deep convolutional approaches for the analysis of Covid-19 using chest X-ray images

**DOI:** 10.1101/2020.05.01.20087254

**Authors:** Joaquim de Moura, Jorge Novo, Marcos Ortega

## Abstract

Covid-19 is a new infectious disease caused by severe acute respiratory syndrome coronavirus 2 (SARS-CoV-2). Given the seriousness of the situation, the World Health Organization declared a global pandemic as the Covid-19 rapidly around the world. Among its applications, chest X-ray images are frequently used for an early diagnostic/screening of Covid-19 disease, given the frequent pulmonary impact in the patients, critical issue to prevent further complications caused by this highly infectious disease.

In this work, we propose complementary fully automatic approaches for the classification of chest X-ray images under the analysis of 3 different categories: Covid-19, pneumonia and healthy cases. Given the similarity between the pathological impact in the lungs between Covid-19 and pneumonia, mainly during the initial stages of both lung diseases, we performed an exhaustive study of differentiation considering different pathological scenarios. To face these classification tasks, we exploited and adapted to this topic a densely convolutional network architecture, which connects each layer to every other layer in a feed-forward fashion. To validate the designed approaches, several representative experiments were performed using images retrieved from different public chest X-ray images datasets. overall, satisfactory results were obtained from the designed experiments, facilitating the doctors’ work and allowing better an early diagnostic/screening and treatment of this relevant pandemic pathology.

## 1 Introduction

A new coronavirus SARS-CoV-2, which causes the disease commonly known as Covid-19, was first identified in Wuhan, Hubei province, China at the end of 2019 [1]. In particular, coronaviruses are a family of viruses known to contain strains capable of causing severe acute infections that typically affect the lower respiratory tract and manifests as pneumonia [2], and therefore, being potentially fatal in humans and a wide variety of animals, including birds and mammals such as camels, cats or bats [3] [4].

On 11 February 2020, the World Health Organization (WHO) declared the outbreak of Covid-19 as a pandemic, noting the more than 118,000 cases of coronavirus disease reported in more than 110 countries, resulting in 4,291 deaths worldwide [5] by that extremely early moment, numbers that were deeply worsened posteriorly. In particular, this disease, among its consequences, also manifests as relevant respiratory disease, being considered a serious public health problem because it can kill healthy adults, as well as older people with underlying health problems or other recognized risk factors, such as heart disease, lung disease, hypertension and diabetes, among others [6]. Furthermore, the Covid-19 virus is transmitted quite efficiently, since an infected person is capable of transmitting the virus to 2 or 3 other people, an exponential rate of increase [7].

Over the years, X-ray examination of the chest plays an important clinical role in detecting or monitoring the progression of different pulmonary diseases, such as emphysema, chronic bronchitis, pulmonary fibrosis, lung cancer or pneumonia, among others [8] [9] [10]. Today, given the severity of the coronavirus pandemic, radiologists are asked to prioritize chest X-rays of patients with suspected Covid-19 infection over any other imaging studies, allowing for more appropriate use of medical resources during the initial screening process and excluding other potential respiratory diseases, a process which is extremely tedious and time-consuming. In this context, the visibility of the Covid-19 infection in the chest X-ray is complicated and requires experience of the clinical expert to analyze and understand the information and differentiate the cases from other diseases of respiratory origin with similar characteristics such as pneumonia, as illustrated in Figure 1. For that reason, a fully automatic system for the classification of chest X-ray images between healthy, pneumonia or specific Covid-19 cases is significantly helpful as it drastically reduces the workload of the clinical staff. Complementary, it may provides a more precise identification of this highly infectious disease, reducing the subjectivity of clinicians in the early screening process, and thus also reducing healthcare costs.

**Figure 1:**
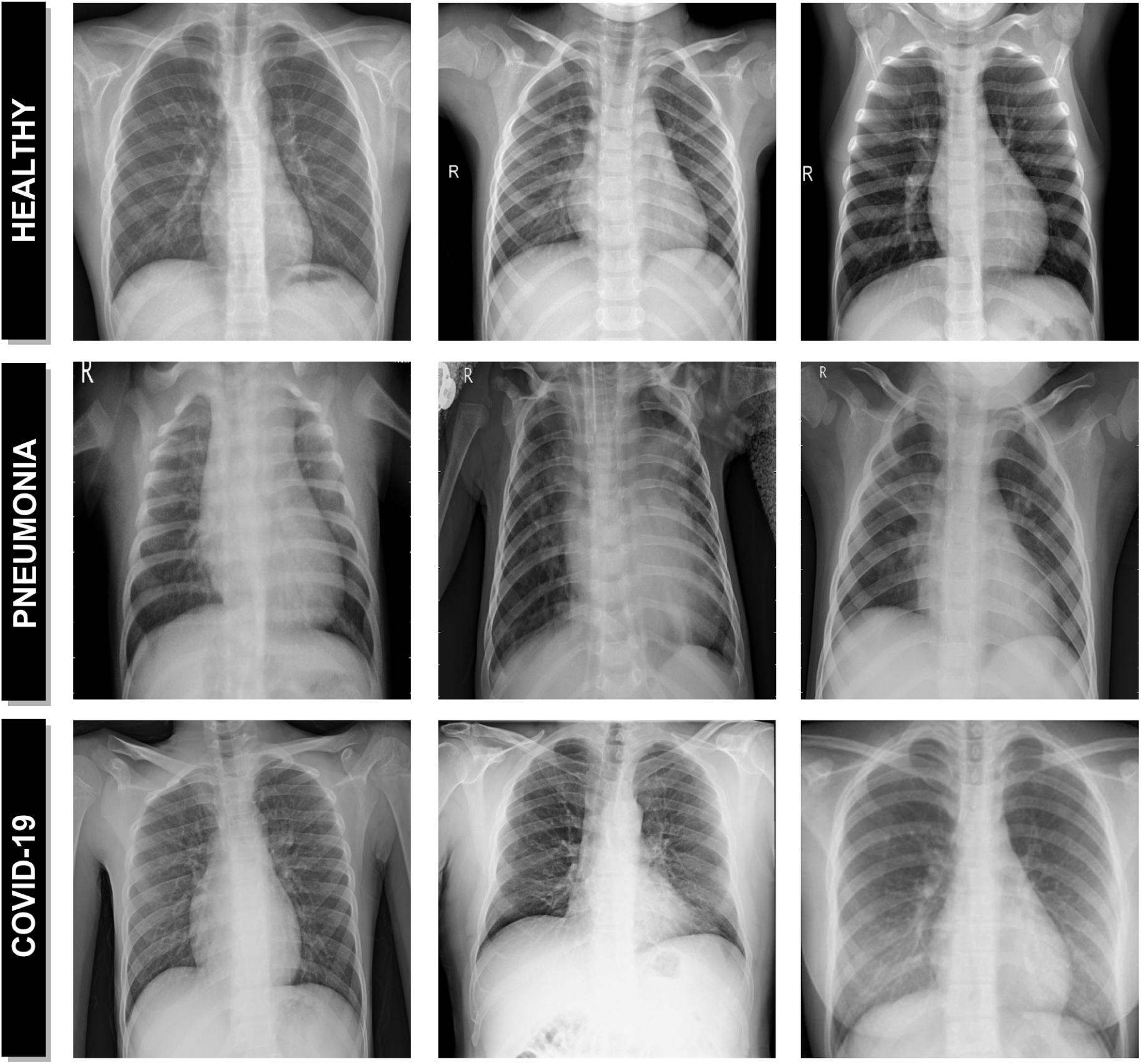
Representative examples of chest X-ray images. 1^st^ row, chest X-ray images from healthy patients. 2^nd^ row, chest X-ray images from patients with pneumonia. 3^rd^ row, chest X-ray images from patients with Covid-19.

Given the relevance of this topic, several approaches were recently proposed using chest X-ray images for the classification of Covid-19. As reference, Sun *et al*. [11] proposed an approach based on deep transfer learning using chest X-ray images for the detection of patients infected with coronavirus pneumonia. In the work of Hassanien *et al*. [12], the authors proposed a methodology for the automatic X-ray Covid-19 lung classification using a multi-level threshold based on Otsu algorithm and support vector machine for the prediction task. Apostolopoulos *et al*. [13] proposed a study on the possible extraction of representative biomarkers of Covid-19 from X-ray images using deep learning strategies. Wang *et al*. [14] proposed a deep convolutional neural network, called COVID-Net, design tailored for the detection of Covid-19 cases from chest radiography images. In the work of Hammoudi *et al*. [15], the authors proposed a deep learning strategy to automatically detect if a chest X-ray image is healthy, pneumonia (bacterial or viral), assuming that a patient infected by the Covid-19, tested during an epidemic period, has a high probability of being a true positive when the result of the classification is a virus.

Despite the satisfactory results obtained by these works, most of them only partially address this recent and relevant problem of global interest, limiting their practical utility for usage and interpretation for support in clinical decision scenarios such as emergency triage for example.

Therefore, in order to offer a more comprehensive methodology we propose in this work complementary fully automatic approaches for the classification of Covid-19, pneumonia and healthy chest X-ray radiographs. To achieve this, we adapted to this issue a densely convolutional network architecture, which generally connects each layer to every other layer in a feed-forward fashion. In this way, the proposed approaches allow to make predictions using complete chest X-ray images of arbitrary sizes, which is very relevant considering the great variability of X-ray devices currently available in health centers. To validate our proposal, exhaustive representative experiments were performed using images compiled from different public image datasets.

The manuscript is organized as follows: Section 2 describes the materials and methods that were used in this research work. Section 3 presents the results and validation of the proposed approaches. Section 4 includes the discussion of the experimental results. Finally, Section 5 presents the conclusions about the proposed systems as well as possible future lines of work in this enormeous topic of interest.

## 2 Materials and Methods

A schematic representation of the proposed paradigm of the different approaches can be seen in Figure 2. The proposed systems receive, as input, a chest X-ray radiography. During the acquisition procedure, the patient is exposed to a small dose of ionizing radiation to produce images of the interior of the chest. The technician will usually be behind a protected wall or in an adjacent room to activate the X-ray machine. The proposed system then uses advanced artificial intelligence techniques to classify chest X-ray images into 3 different clinical categories: healthy, pneumonia or Covid-19. As a result, the system provides useful clinical information for the initial screening process and for subsequent clinical analyses.

**Figure 2:**
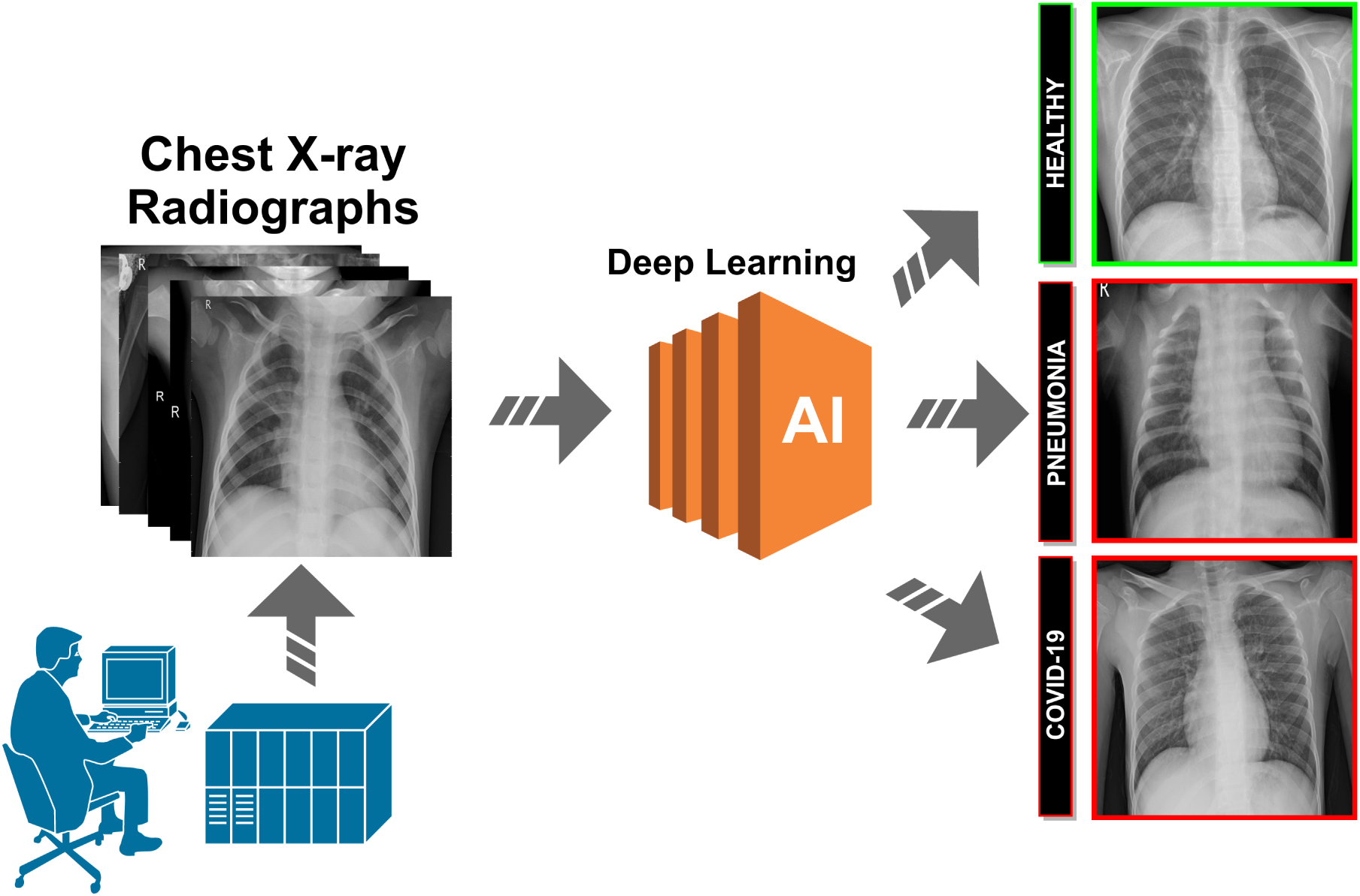
Representative scheme of the proposed methodology.

### 2.1 Computational approaches for pathological X-ray image classification

Given the similarity between the pathological impact in the lungs between Covid-19 and common types of pneumonia, mainly during the initial stages of both lung diseases, we performed an exhaustive analysis of differentiation considering different pathological scenarios. In this line, we proposed 4 different and independent computational approaches for the classification of Covid-19, pneumonia and healthy chest X-ray radiographs. Each of these approaches is explained in more detail below:

#### 2.1.1 1^*st*^ approach. Healthy vs Pneumonia, tested with Covid-19

Using as reference a consolidated public image dataset for the identification of pneumonia subjects with a considerable amount of image samples [16], we firstly used, as baseline, a trained model for the differentiation of healthy and pneumonia chest X-ray images taking advantage of this large amount of available information. Subsequently, we tested the potential of this approach to classify chest X-ray radiographs of patients diagnosed with Covid-19 and measure their similarity with both situations. In this way, we can analyze the percentage of chest X-ray images of patients with Covid-19 that may be classified as pneumonia, giver their relation in the pathological pulmonary impact.

#### 2.1.2 *2^nd^* approach, Healthy vs Pneumonia/Covid-19

Given the pathological similarity between pneumonia and covid-19 subjects, subsequently we designed an screening process that analyzes the degree of separability between healthy and pathological chest X-ray radiographs, considering these both pathological scenarios. In this sense, we include the chest X-ray images of patients diagnosed with pneumonia or Covid-19 under the same class in a training of the model to predict 2 different categories: pathological and healthy cases.

#### 2.1.3 3*^rd^* approach, Healthy/Pneumonia vs Covid-19

Additionally, we adapted and trained a model to specifically identify Covid-19 subjects, measuring the capability of differentiation not only from healthy subjects, but also from those pathological cases with a significant similarity, as patients suffering from pneumonia. With this in mind, our system was designed to identify two different classes, including healthy and patients with pneumonia in the same category.

#### 2.1.4 4*^th^* approach, Healthy vs Pneumonia vs Covid-19

Finally, we designed another approach to simultaneously determine the degree of separability between the 3 categories of chest X-ray images considered in this work. To this end, we trained a model with a set of chest X-ray radiographs of the 3 different classes: healthy subjects, patients diagnosed with pneumonia, patients diagnosed with Covid-19.

### 2.2 Network Architecture

The use of deep learning architectures has been rapidly increasing in the field of medical imaging, including computer-aided diagnosis system and medical image analysis for more accurate screening, diagnosis, prognosis and treatment of many relevant diseases [17] [18]. These architectures have proven to be superior in both accuracy and predictive efficiency compared to classical machine learning techniques [19]. In this work, we use a densely convolutional network architecture inspired by DenseNet proposed by Huang *et al*. [20], given its simplicity and potential, providing adequate results for many similar classification tasks in pulmonary diseases [21] [22] [23]. In our case, this dense network architecture was initialized with weights from a model pretrained on ImageNet, connecting each layer to every other layer in a feed-forward fashion within each dense block. On the one hand, we benefit from the pre-trained model weights making the learning process much more efficient, in other words, the model converges fast because his weights are already stabilized initially. On the other hand, it significantly reduces the amount of labeled data required for model training. For each layer, feature maps from all preceding layers are processed as separate inputs, while their own feature maps are transferred as inputs to all subsequent layers. In this study, we employed a modification of the original structure of the DenseNet-161 architecture, as illustrated in Figure 3. In particular, we have adapted the classification layer of the architecture used to support the output according to the specific requirements of each proposed approach, which is to categorize chest X-ray images into 2 or 3 different clinical classes considering healthy, pneumonia and Covid-19.

**Figure 3:**
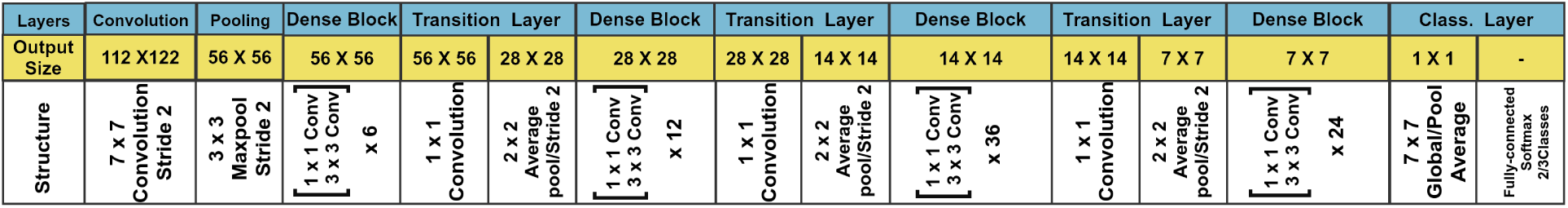
An illustration of the DenseNet architecture that was adapted for the different and independent computational approaches of this work.

### 2.3 Training

Regarding the training stage of the different approaches, considering the limited amount of covid-19 subjects, we decided that the employed chest X-ray radiographs dataset was randomly divided into 3 smaller datasets, specifically with 60% of the cases for training, 20% for validation and the remaining 20% for testing. Additionally, the classification step was performed with 5 repetitions, being calculated the mean cross-entropy loss [24] and the mean accuracy to illustrate the general performance and stability of the proposed approaches. The DenseNet-161 architecture was trained using Stochastic Gradient Descent (SGD) with a constant learning rate of 0.01, a mini-batch size of 4 and a first-order momentum of 0.9. In particular, SGD is a simple but highly efficient approach for the discriminative learning of classifiers under convex loss functions [25].

### 2.4 Data Augmentation

Data augmentation is a widely used strategy that enables practitioners to significantly increase the diversity of data available for training models, reducing the overfitting and making the models more robust [26] [27]. This is especially significant in our case, given the limited amount of positive Covid-19 cases that were used in the different approaches. To this end, we applied different configurations of affine transformations to increase the training data and improve the performance of the neural network architecture that was used to classify chest X-rays images. In particular, we automatically generate additional training samples through a combination of scaling with horizontal flipping operations, considering the common variety of possible resolutions as well as the simmetry of the human body.

### 2.5 Dataset

The training and evaluation of the deep convolutional models were performed with chest X-ray radiographs that were taken from 3 different chest X-ray public datasets of reference: the Chest X-ray (Pneumonia) dataset [16], the (Covid-19) Image Data Collection dataset [28] and the (Covid-19) SIRM dataset [29].

The Chest X-ray (Pneumonia) dataset of the Radiological Society of North America (RSNA)[16] is composed by of 5,863 chest X-ray radiographs. This public dataset was labeled into 2 main categories: healthy patients and patients with different types of pneumonia (viral and bacterial) presenting, therefore, a high level of heterogeneity.

Currently, public chest X-ray datasets of patients diagnosed with Covid-19 are very limited. Despite this important restriction, we have built a dataset composed of 207 radiographs, 155 were taken from (Covid-19) Image Data Collection dataset [28] and 52 were taken from the (Covid-19) SIRM dataset of the Italian Society of Medical Radiology [29].

### 2.6 Evaluation

In order to test its suitability, the designed paradigm of the different approaches was validated using different statistical metrics commonly used in the literature to measure the performance of computational proposals in similar medical imaging tasks. Accordingly, Precision, Recall, F1-score and Accuracy were calculated for the quantitative validation of the classification results. In particular the first three metrics are calculated for each one of the considered classes in the different experiments are they are more meaningful in that way.

Mathematically, these statistical metrics are formulated as indicated in Equations 1, 2, 3 and 4, respectively. These performance measures use as reference the True Negatives (TN), False Negatives (FN), True Positives (TP) and False Positives (FP):

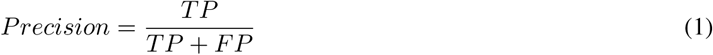

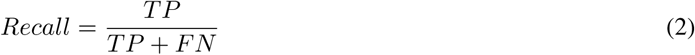

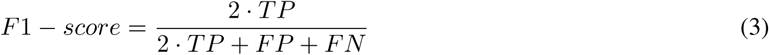

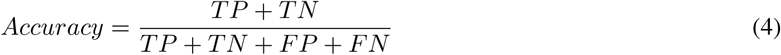

## 3 Experimental Results

To evaluate the suitability of the different proposed approaches in the pathological classification related to Covid-19 in chest X-ray images, we conducted different complementary experiments, taking as reference the available datasets. In particular, for each experiment, we performed 5 independent repetitions, each time with a different random selection of the samples splits, specifically with 60% of the cases for training, 20% for validation and the remaining 20% for testing. Additionally, the training stage was stopped after 200 epochs given the lack of significant further improvement in both accuracy and cross-entropy loss results.

### 3.1 1*^st^* experiment: Healthy vs Pneumonia, tested with Covid-19

Given the availability of a public image dataset of reference with a significant number of healthy and pneumonia chest X-ray images [16], we trained a model to obtain a consolidated approach to distinguish between healthy patients and different pathological cases of pneumonia.

In this line, we designed an experiment with a total of 5,856 X-ray images, being 1,583 from healthy patients and 4,273 from patients with pneumonia. Figure 4 shows the performance that was obtained using the deep learning architecture after, as previously indicated, 5 independent repetitions in the training and validation stages.

**Figure 4:**
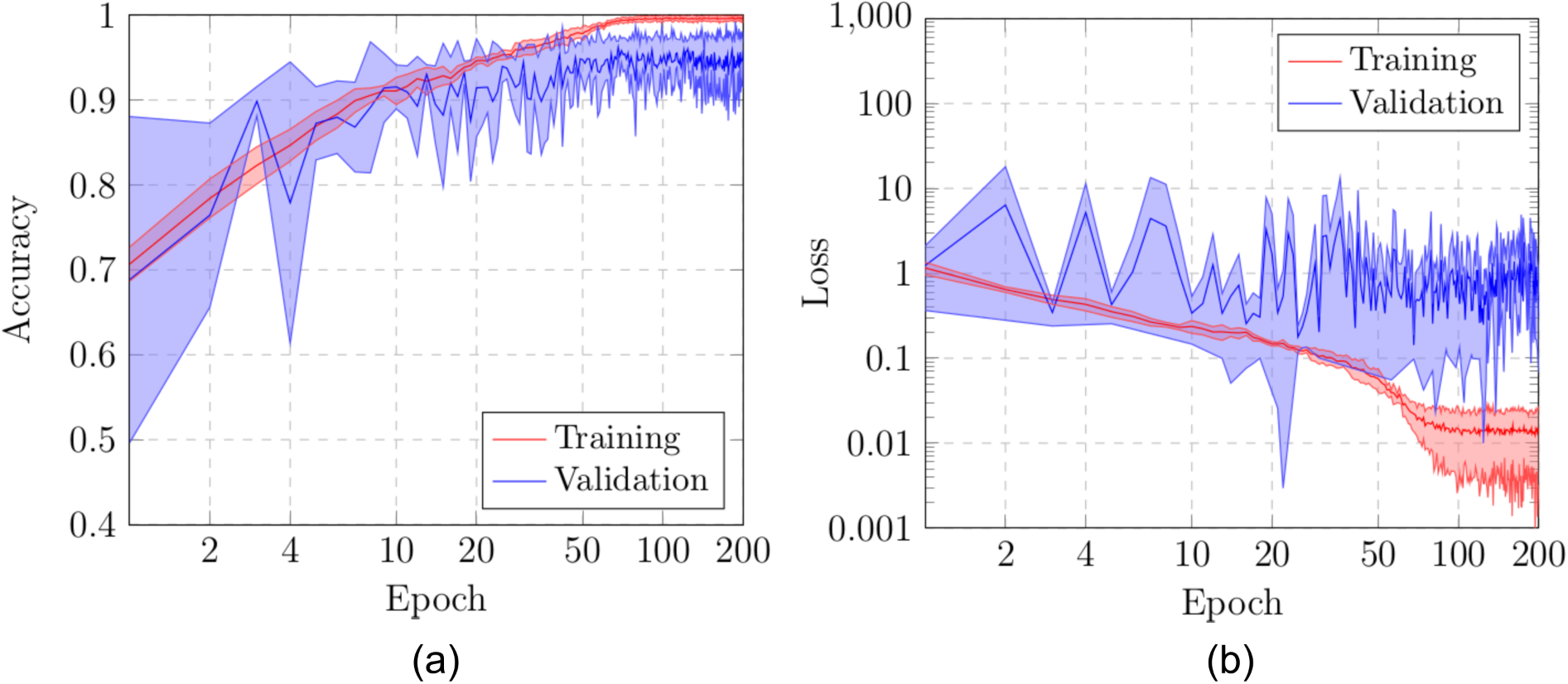
Results of the first experiment after 5 independent repetitions. (a) Mean ± standard deviation training accuracy. (b) Mean ± standard deviation training loss. In both cases, a logarithmic scale has been set to correctly display the values for a better understanding of the results.

The method achieved satisfactory results, reaching a best average accuracy of 0.9971 ± 0.0026 for training in epoch 194 and the best average accuracy for validation of 0.9624 ± 0.0067 in epoch 68, as we can see in Figure 4(a)). Furthermore, in Figure 4(b) we can observe that the model converged quite fast in the training and validation steps in terms of the loss cross-entropy function.

In Table 1, we can see the precision, recall and F1-score results obtained at the test stage, providing an accuracy of 0.97. All the results obtained show the robustness of the proposed system in the classification of the different pathological cases of pneumonia and healthy patients.

**Table 1:** Precision, recall and F1-score results obtained at the test stage for the classification of chest X-ray images between Healthy vs Pneumonia cases.

| Cases | Precision | Recall | F1-score |
| --- | --- | --- | --- |
| Healthy | 0.95 | 0.93 | 0.94 |
| Pneumonia | 0.98 | 0.98 | 0.98 |

In addition to measure the capability of the adapted architecture to this pathological analysis, another relevant goal of this experimentation was to conduct a comprehensive analysis about the potential similarity of the Covid-19 subjects with pneumonia scenarios, that is, the percentage of chest X-ray images of patients with Covid-19 that are classified as pneumonia with the previously trained network.

Thus, we extended the experiments of this case using an additional blind test dataset. In particular, this dataset consisted of the used 207 X-ray images of patients diagnosed with Covid-19. As we can see in Table 2, the proposed system achieves satisfactory results in terms of precision, recall and F1-score, considering Covid-19 cases correctly classified as pneumonia in opposition to healthy cases. In this scenario, the system achieves an accuracy of 0.83, demonstrating that the system is capable of correctly screening X-ray images of Covid-19 patients in the pathological pneumonia category.

**Table 2:** Precision, recall and F1-score results obtained at the test stage for the classification of chest X-ray images from Covid-19 patients between Healthy vs Pneumonia cases.

| Cases | Precision | Recall | F1-score |
| --- | --- | --- | --- |
| Healthy | — | — | — |
| Pneumonia | 1.00 | 0.83 | 0.90 |

### 3.2 *2^nd^* experiment: Healthy vs Pneumonia/Covid-19

Under the results of the analysis of the first approach, we designed another scenario with a screening context including Covid-19 cases by separating the pathological pneumonia and Covid-19 subjects with respect to healthy images

With this in mind, in this case, the designed experiment included a total of 1242 X-ray images, being 828 from healthy patients whereas 207+207 from Covid-19 and pneumonia patients, respectively. In this case, we randomly selected the 828 healthy images and the 207 pneumonia images from the total amount of the used image dataset [16]. Having the limiting factor of 207 Covid-19 images, we balanced the amount of the other cases to obtain a proportion of 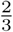 and 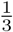 between the negative and positive classes.

Figure 5 shows the performance that was obtained from the training and validation stages using the proposed dataset through 5 independent repetitions. As we can see in Figure 5(a), the best mean accuracy that was produced is 0.9989 ± 0.0011 for training in epoch 121 and the best average accuracy for validation of 0.9887 ± 0.0044 in epoch 167. In addition, the proposed approach achieved its stability in the loss cross-entropy function both for training and for validation after epoch 100, as we can see in Figure 5(b)).

**Figure 5:**
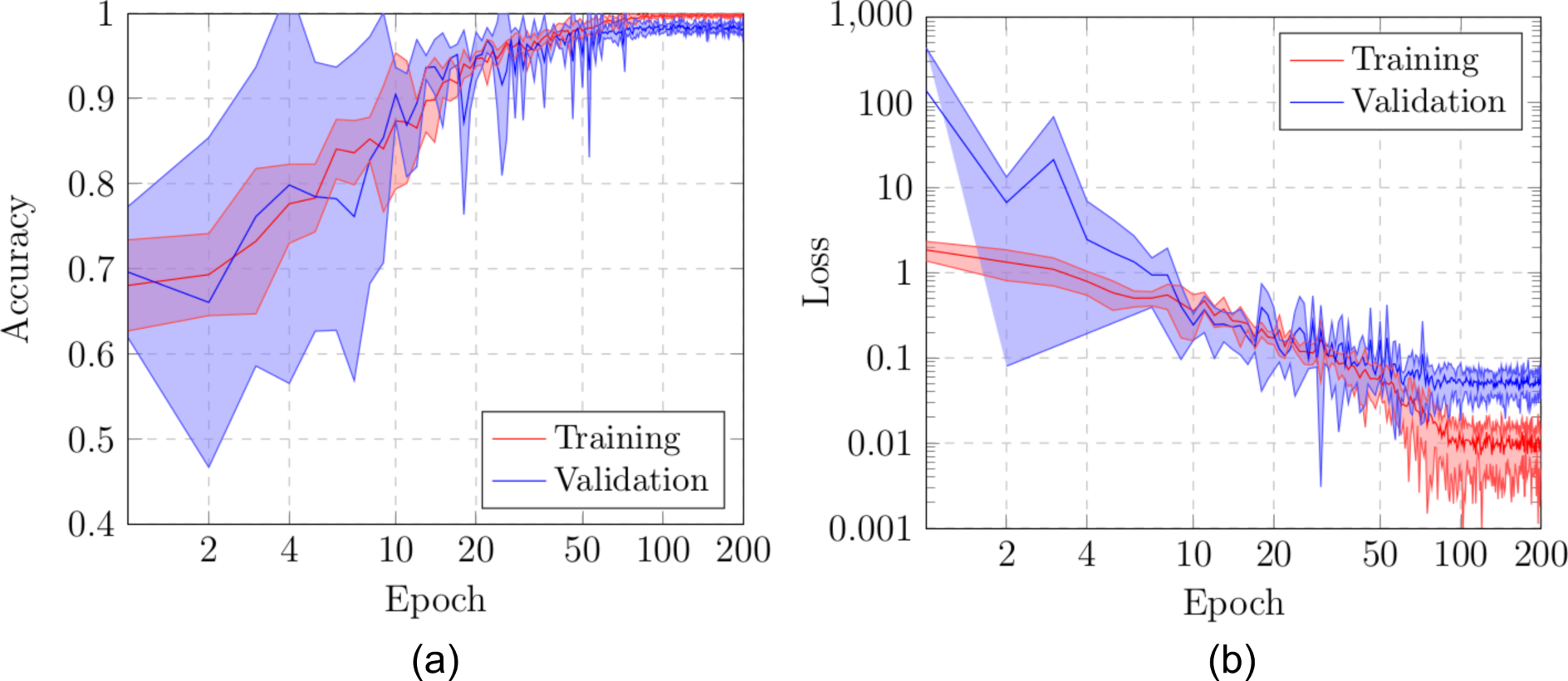
Results of the second experiment after 5 independent repetitions. (a) Mean ± standard deviation training accuracy. (b) Mean ± standard deviation training loss. In both cases, a logarithmic scale has been set to correctly display the values for a better understanding of the results.

Results in Table 3 show the performance measurements obtained in the test stage, in terms of precision, recall and F1-score for each class. As we can see, satisfactory results were provided as a global accuracy of 0.98 for both categories. Thus, the results obtained show that this screening approach is capable of successfully separating the considered pathological cases from the healthy ones.

**Table 3:**
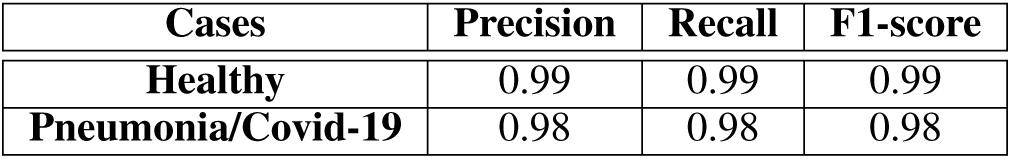
Precision, recall and F1-score results obtained at the test stage for the classification of chest X-ray images between Healthy vs Pneumonia/Covid-19 cases.

| Cases | Precision | Recall | F1-score |
| --- | --- | --- | --- |
| Healthy | 0.99 | 0.99 | 0.99 |
| Pneumonia/Covid-19 | 0.98 | 0.98 | 0.98 |

### 3.3 3*^rd^* experiment: Healthy/Pneumonia vs Covid-19

The third scenario was designed to evaluate the performance of the proposed approach to specifically distinguish between cases of patients with Covid-19 from other similar cases such as pneumonia or also from healthy patients. Thus, we measure the potential separability between those potentially similar pathological cases of Covid-19 from pneumonia.

To do so, we used a total of 621 X-ray images, being 207 from Covid-19, 207 from patients with pneumonia and 207 from healthy patients. Once again, those subjects from pneumonia and healthy patients were randomly selected to obtain a proportion of 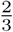 and 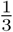 between negative and positive classes, respectively. Figure 6 shows the performance that was obtained from the DenseNet-161 architecture after 5 independent repetitions in both the training and validation steps. In particular, as we can observe in Figure 6(a), our model reaches high stability after 75 epochs, with a better average accuracy of 0.9973 ± 0.0031 for training in epoch 153 and a better average accuracy for validation of 0.9741 ± 0.0067 in epoch 111. Complementary, in Figure 6(b) we can observe a similar behavior with the cross-entropy loss function,

**Figure 6:**
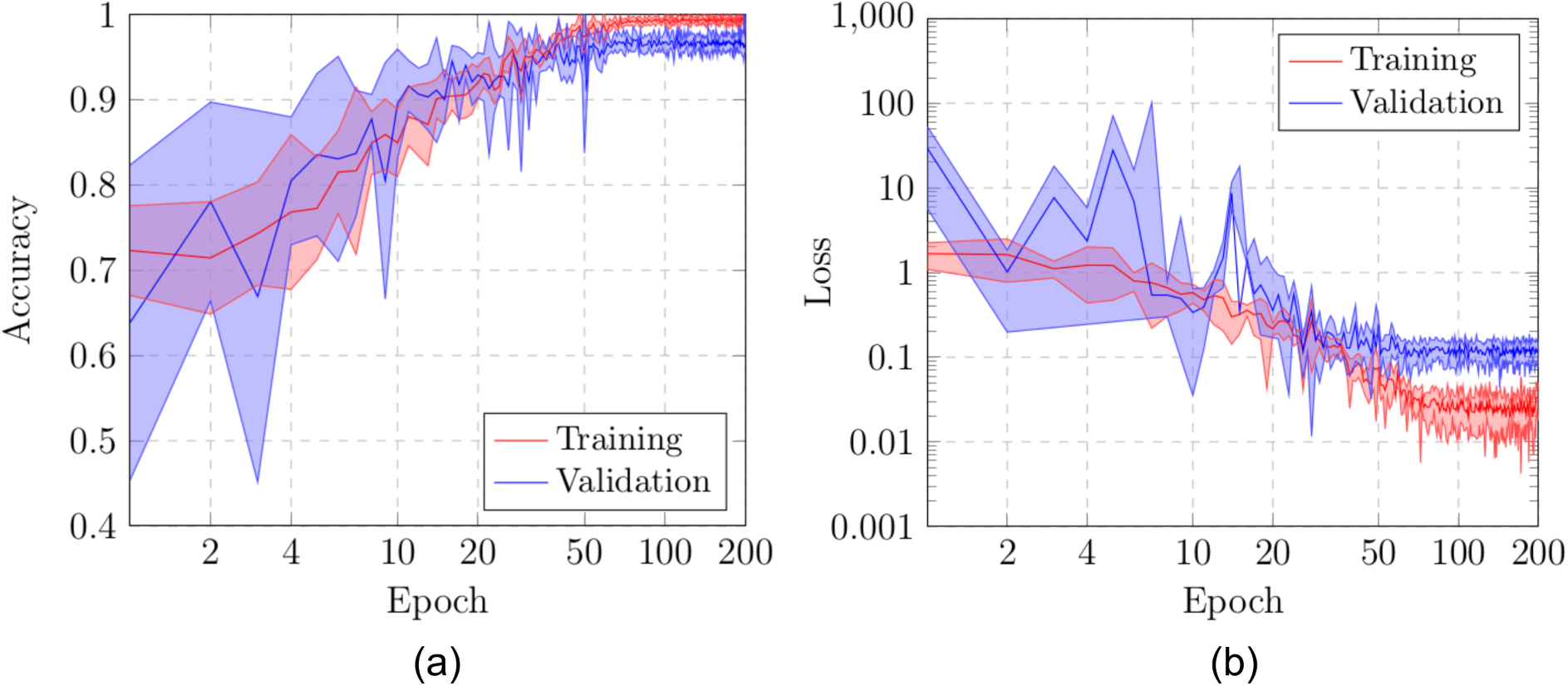
Results of the third experiment after 5 independent repetitions. (a) Mean ± standard deviation training accuracy. (b) Mean ± standard deviation training loss. In both cases, a logarithmic scale has been set to correctly display the values for a better understanding of the results.

In Table 4 we present the quantitative performance of the proposed system in the test dataset, in terms of precision, recall and F1-score. Our method shows a good performance for both categories, providing a global accuracy of 0.98 demonstrating the robustness of the proposed system to distinguish between cases of patients with Covid-19 from other cases such as pneumonia or health. This result is specially significant considering the separation of similar pathological scenarios as pneumonia and Covid-19.

**Table 4:** Precision, recall and F1-score obtained at the test stage for the classification of chest X-ray images between Healthy/Pneumonia vs Covid-19 cases.

| Cases | Precision | Recall | F1-score |
| --- | --- | --- | --- |
| **Healthy/Pneumonia** | 0.96 | 1.00 | 0.98 |
| **Covid-19** | 1.00 | 0.93 | 0.97 |

### 3.4 *4^th^* experiment: Healthy vs Pneumonia vs Covid-19

In this last scenario, considering the satisfactory results of the other considered approaches, we also trained another model to directly analyze the performance of the proposed system to separate the chest X-ray radiographs into 3 different categories: healthy, pneumonia and Covid-19. To do so, we designed a complete experiment using a total of 621 X-ray radiographs, being 207 from Covid-19, 207 from patients with pneumonia and 207 from healthy patients.

In this case, again, we randomly selected the healthy and pneumonia images to balance the amount of available images for the 3 considered classes. Figure 7 shows the performance of the proposed system using training and validation sets after 5 independent repetitions. As we can see in Figure 7(a), the best mean accuracy that was produced is 0.9967 ± 0.0029 for the training stage in epoch 166 and the best average accuracy for validation of 0.9725 ± 0.0185 in epoch 189. In the same line, the proposed system achieved its stability in the loss cross-entropy function both for training and for validation after epoch 100, as we can see in Figure 7(b).

**Figure 7:**
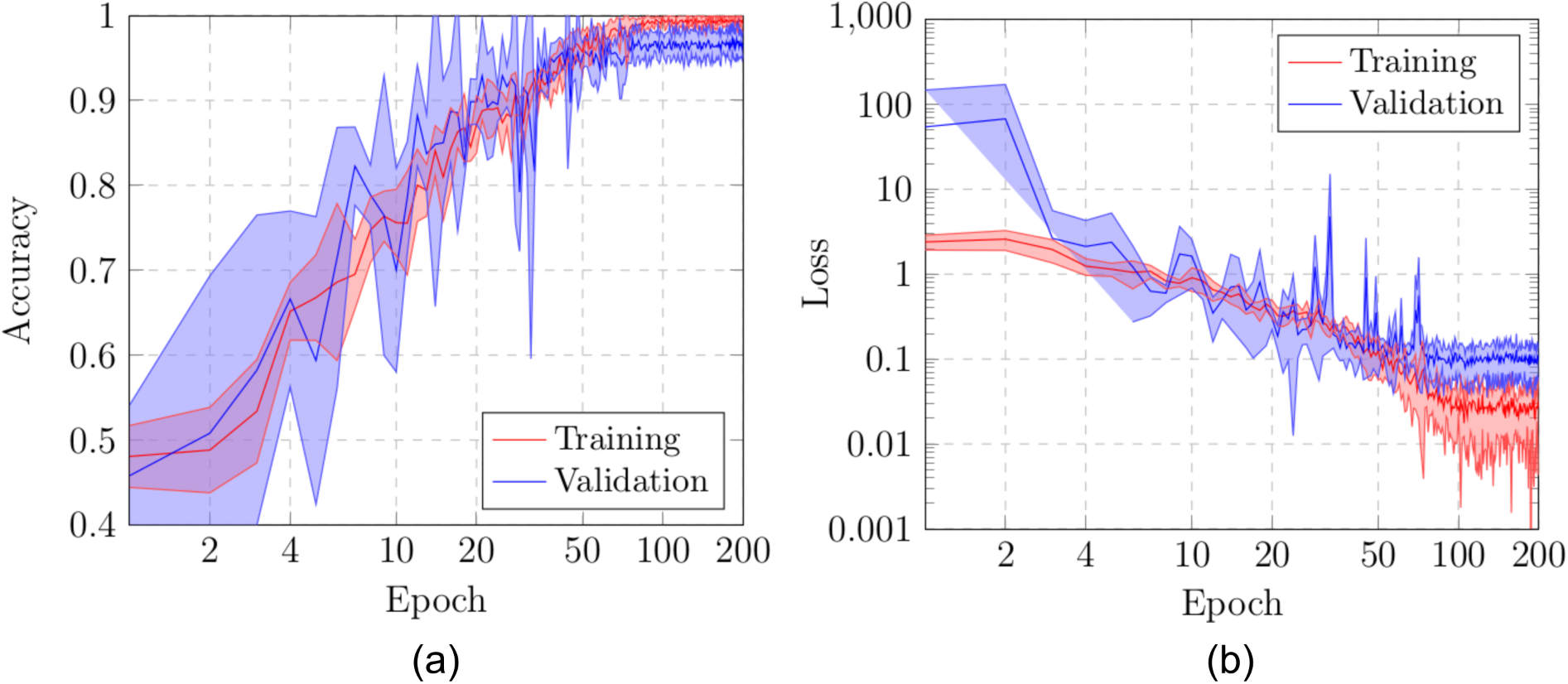
Results of the fourth experiment after 5 independent repetitions. (a) Mean ± standard deviation training accuracy. (b) Mean ± standard deviation training loss. In both cases, a logarithmic scale has been set to correctly display the values for a better understanding of the results.

In Table 4, we can see the precision, recall and F1-score results obtained at the test stage, providing an accuracy of 0.99. As we can see, the trained model is capable to predict with accuracy of 0.99 all the mentioned categories. Generally, the obtained results in all the cases are satisfactory, demonstrating the robustness of the proposed system in the classification of the 3 categories of chest X-ray images considered in this work.

**Table 5:** Precision, recall and F1-score results obtained at the test stage for the classification of chest X-ray images between Healthy vs Pneumonia vs Covid-19 cases.

| Cases | Precision | Recall | F1-score |
| --- | --- | --- | --- |
| **Healthy** | 0.98 | 1.00 | 0.99 |
| **Pneumonia** | 1.00 | 1.00 | 1.00 |
| **Covid-19** | 1.00 | 0.98 | 0.99 |

## 4 Discussion

In this work, we analyzed different and complementary fully automatic approaches for the classification of healthy, pneumonia and Covid-19 chest X-ray radiographs. All the experimental results demonstrate that the proposed system is capable of successfully distinguishing healthy patients from different pathological cases of pneumonia and Covid-19. This pathological differentiation is understandable given the abnormality of the pathological scenarios with respect to normal patients and also the pathological impact in the lungs of both pneumonia and Covid-19 diseases. But also, it is significant the accurate capability of differentiation also of Covid-19 patients from other with pneumonia, which were correctly separated in the proposed third and forth approaches, also corroborated by the experiments with the first approach. In addition, the proposed system allows to make accurate predictions using chest X-ray images of arbitrary sizes, which is very relevant considering the great variability of X-ray devices currently available in the healthcare centers.

Despite the attained satisfactory performance, the proposed system obtain an acceptably small number of misclassified cases. In particular, some misclassification is caused by the poor contrast of the X-ray images used in this work. Other times, in some cases, there is a great similarity between Covid-19 and pneumonia, mainly during the initial stages of both diseases. In figure 8, we can see representative examples illustrating the significant variability of the possible scenarios that are represented in this research work.

**Figure 8:**
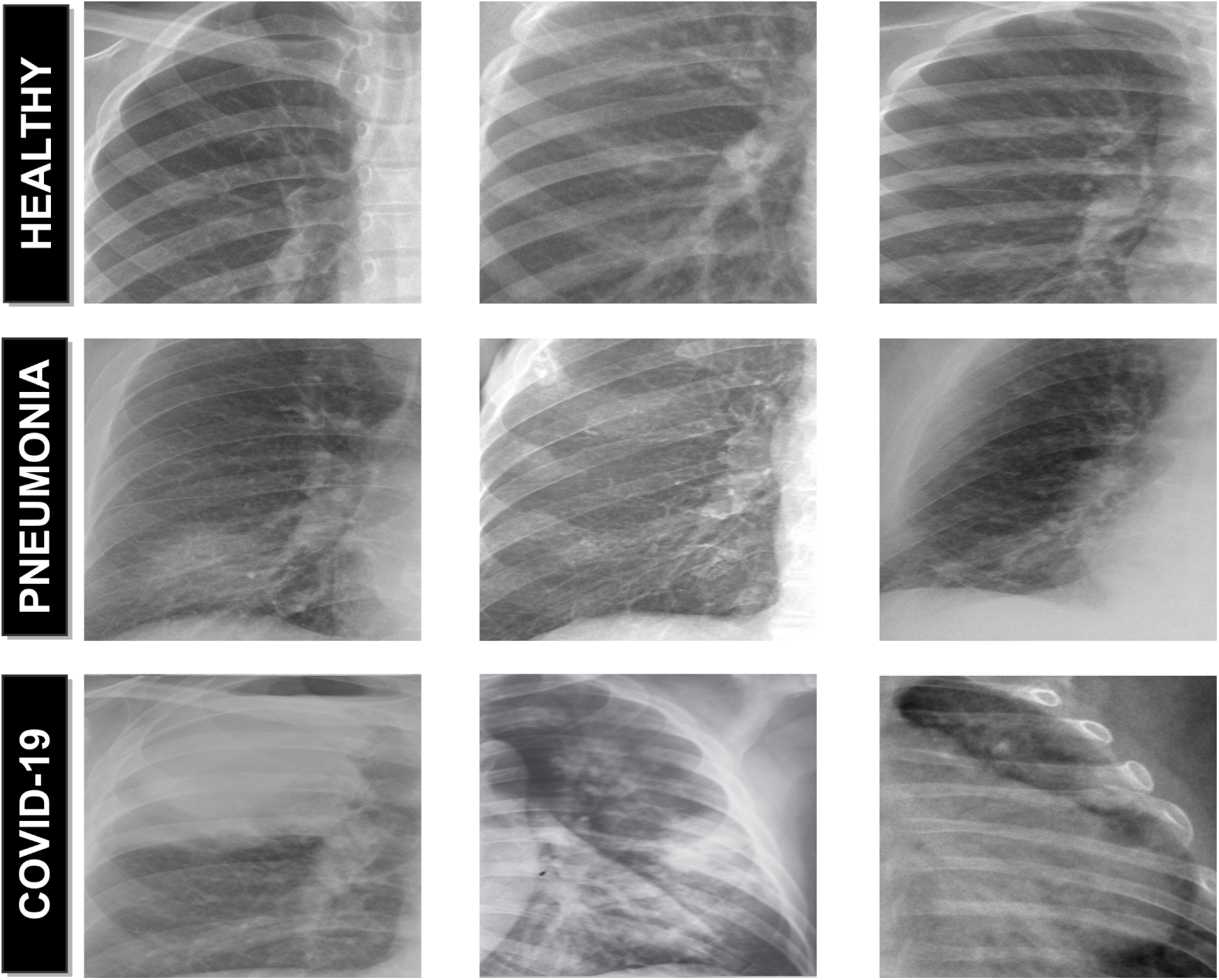
Representative examples of lung regions. 1*^st^* row, lung healthy regions. 2*^nd^* row, lung regions affected by the pneumonia disease. 3*^rd^* row, lung regions affected by the Covid-19 disease.

As no exhaustive classification method for X-ray Covid-19 images has been published to date, we cannot make any comparison with other state-of-the-art approaches. Instead, we use different public datasets for the evaluation of our proposed system, validating their accuracy in comparison with manual annotations from different clinical experts.

## 5 Conclusions

Coronaviruses are a large family of viruses that can cause disease in both animals and humans. In particular, the new coronavirus SARS-CoV-2, also know Covid-19, was firstly detected in December 2019 in Wuhan City, Hubei Province, China. Given its drastic spread, the WHO declared a global pandemic as the Covid-19 rapidly spreads across the world. In this context, chest X-ray images are widely used for early screening by clinical experts, allowing for more appropriate use of other medical resources during initial screening.

In this work, we proposed complementary fully automatic approaches for the classification of Covid-19, pneumonia and healthy chest X-ray radiographs. To achieve this, we adapted a densely convolutional network architecture, which connects each layer to every other layer in a feed-forward fashion. We evaluated the robustness and accuracy of the different classification approaches, obtaining satisfactory results for all the experiments that were proposed using different public image datasets of reference. Despite the complex and challenging scenario, the proposed approaches has proven to be robust and reliable, facilitating a more complete and precise analysis of the pathological lung regions and, consequently, the production of more adjusted treatments of this highly infectious disease.

As future work, we plan to expand the proposed methodology with the incorporation of other relevant lung diseases, such as chronic bronchitis, emphysema, or lung cancer. Additionally, further analysis with larger and maybe more comprehensive chest X-ray datasets should be done in order to reinforce the conclusions of this work.

## Data Availability

not aplicable

## Acknowledgement

This work is supported by the Instituto de Salud Carlos III, Government of Spain and FEDER funds of the European Union through the DTS18/00136 research projects and by the Ministerio de Ciencia, Innovación y Universidades, Government of Spain through the RTI2018-095894-B-I00 research projects. Also, this work has received financial support from the European Union (European Regional Development Fund - ERDF) and the Xunta de Galicia, Centro de Investigation del Sistema universitario de Galicia, Ref. ED431G 2019/01.

